# Systemic and organ-specific proteomic aging clocks as predictors of cancer incidence and survival: insights from the Finnish Twin Cohort

**DOI:** 10.64898/2026.10.14.26365410

**Authors:** V Lukander, J Kaprio, X Wang, M Ollikainen, G Drouard

**Author notes:** **Correspondence to:** Volter Lukander, Institute for Molecular Medicine Finland, FIMM, University of Helsinki P.O. Box 20, FI-00014 Helsinki, Finland. **Novelty and Impact:** This study utilizes large-scale plasma proteomics within a longitudinal twin cohort to evaluate how systemic and organ-specific proteomic aging profiles relate to cancer incidence and mortality. We demonstrate that while systemic proteomic aging strongly predicts all-cause mortality, the organ-specific Immune clock stands out as a robust, independent predictor of cancer lethality. These findings highlight the potential of proteomic profiling for targeted cancer risk stratification.

## Abstract

Despite recent evidence of the ability of systemic and organ-specific proteomic aging clocks to predict a large range of age-related diseases, their utility in cancer research remains largely underexplored. In this study, we evaluated the predictive ability of systemic and organ-specific proteomic aging clocks for (i) all-cause mortality, (ii) cancer-related mortality, and (iii) cancer incidence in a sample of Finnish twins (N = 401, mean age at baseline 62.3 years) with Olink proteomic data. Participants were followed for a mean of 10.6 years (SD ± 1.6), with estimates from four systemic and 18 organ-specific proteomic aging clocks calculated at baseline. Cox proportional hazards regression models adjusted for sex and age at baseline identified 15 proteomic aging clocks as significant predictors of all-cause mortality (N_cases_ = 28) after multiple testing correction (FDR < 0.05), with 11 remaining significant after lifestyle adjustment (N_cases_ = 27). For several clocks, effect sizes were large, with hazard ratios (HR) approximately doubling per one standard deviation (SD) increase in clock estimates. In predicting cancer-related deaths among participants without a prior cancer diagnosis at baseline (N_cases_ = 9), only the Immune clock showed a significant positive association after multiple testing correction in basic (HR = 2.94, 95% CI: 1.88–4.59) and lifestyle-adjusted models (HR = 3.31, 95% CI: 1.78–6.17). No associations with cancer incidence (N_cases_ = 34 basic, N_cases_ = 31 adjusted) survived multiple testing correction, although several nominal associations were observed. In conclusion, our findings suggest that proteomic aging clocks are strong predictors of all-cause and cancer-specific mortality, independent of lifestyle factors. Proteomic aging clocks may further serve as valuable tools for stratifying cancers by mortality risk.

## Introduction

Cancer is one of the leading causes of death worldwide, accounting for nearly 10 million deaths annually. While cancer-specific mortality is primarily dictated by stage at diagnosis, tumor biology, and host frailty, cancer incidence is strongly predicted by chronological age and cumulative lifestyle exposures(1–3). Unlike chronological age, biological age reflects an individual’s physiological decline by integrating the cumulative impact of genetics and environmental factors. Consequently, it may predict cancer risk and mortality better than chronological age alone, making molecular aging models, referred to as aging clocks, a powerful framework for studying age-related diseases.

Early aging clocks were primarily developed using DNA methylation (DNAm) data, such as Horvath or GrimAge clocks, capitalizing on the discovery that specific methylation patterns change predictably and in a clock-like manner with advancing age(4,5). Biological age acceleration, defined as a higher algorithmic age relative to chronological age, has since been widely used as a predictor for chronic disease risk(6). In cancer epidemiology, epigenetic age acceleration is significantly associated with both an increased risk of developing tumors and subsequent mortality(7). For instance, accelerated GrimAge and PhenoAge have been linked to higher breast(8) and lung cancer risk(9). Furthermore, longitudinal studies in the Finnish Twin Cohort (FTC) have demonstrated that DNAm age acceleration can increase as a cancer diagnosis approaches, though its long-term predictive power varies(10). Thus, studies utilizing epigenetic aging clocks have successfully demonstrated that molecular measures of biological aging can predict cancer outcomes, paving the way for exploring other omics-based aging biomarkers.

More recently, the availability of large-scale proteomic data from approximately fifty thousand participants in the UK Biobank, generated as part of the UK Biobank Pharma Proteomics Project (UKB-PPP), has greatly expanded the development of proteomic aging clocks(11). These clocks show promise in capturing interindividual variability in chronic disease onset and development(12). However, the potential of proteomic aging clocks for cancer research remains insufficiently characterized, largely due to their recent development and the relatively recent release of UKB-PPP data to the research community. In parallel, methodological refinements have led to the development of so-called organ-specific aging clocks(13), which aim to estimate biological aging at the level of individual tissues or organs rather than providing a single systemic measure(14). This approach is motivated by the core principle that aging processes may not occur uniformly across tissues and organs(13). As with systemic proteomic aging clocks, the extent to which organ-specific proteomic aging clocks may provide insights relevant to cancer research remains to be explored.

Using a sample of Finnish twins linked to national cancer registries and with available Olink proteomic data, we evaluated the predictive ability of a panel of systemic and organ-specific proteomic aging clocks for all-cause mortality, cancer-related mortality, and cancer incidence. We show that proteomic aging clocks enable identification of individuals at elevated risk of death from cancer.

## Methods

### Finnish Twin Cohort

This study examined a subset of twins from the older FTC, referred to as the Essential Hypertension Epigenetic study (EH-Epi)(15), which includes twin pairs with varying differences in blood pressure between the co-twins. Twin pairs were recruited through a questionnaire mailed to them in 2011 and later invited to an in-person clinical visit between 2012 and 2014 during which the twins were interviewed and provided blood samples. This clinical visit is defined as the start of follow-up in the present study. Olink proteomic data and derived proteomic aging clock estimates were later generated from the plasma samples of 401 of these twins, as described in more detail below. The FTC data used in the analyses are deposited in the Biobank of the Finnish Institute for Health and Welfare (https://thl.fi/en/our-services/thl-biobank1).

### Cancer incidence and survival

Cancer history, incident diagnoses, and causes of death were obtained through linkage to the Finnish Cancer Registry and national health registries under Findata permit THL/6353/14.06.00/2023. All-cause mortality was evaluated across the full cohort (N = 401), during which 28 deaths occurred (27 deaths among 384 participants with complete lifestyle covariates). Participants with a diagnosed malignancy prior to baseline (n = 46) were excluded from cancer-specific analyses. In cancer-specific mortality analyses of the remaining 355 cancer-free participants, 9 cancer-related deaths were recorded (6 males, 3 females). Lung cancer was the leading cause of cancer death (n = 3), all other fatal cancers were of unique primary sites. Non-cancer deaths were censored at the date of death. Over the same follow-up period, 34 incident cancer cases were diagnosed in the cancer-free cohort (18 males, 16 females; 9 prostate, 7 breast, 4 skin cancers including melanoma but excluding basal cell carcinoma, 3 lung, and 11 other sites). Following restriction to participants with complete lifestyle covariate data (N = 342), 9 cancer deaths and 31 incident cancer cases remained.

### Proteomic data processing & proteomic aging clocks

Proteomic data were generated as part of the FinnGen study(16) for N=401 twins from blood plasma samples using the Olink platform (Olink Explore 3072 version 3.0.5), which employs antibody-based technology via proximity extension assay (Olink Proteomics AB, Uppsala, Sweden). A full description of the dataset is available elsewhere(17). Data processing included exclusion of sample outliers and verification of Olink’s internal quality control criteria.

We calculated a total of 22 systemic and organ-specific proteomic aging clocks, trained either to predict chronological age (first-generation), mortality (second-generation), or healthspan (third-generation) in UK Biobank participants. Systemic clocks included the ProtAge (first-generation)(12), Proteomic Aging Clock (PAC; second-generation)(18), Conventional systemic clock (second-generation)(19), and Healthspan Proteomic Score (HPS; third-generation)(20). Detailed calculations of these clocks have been provided elsewhere(21).

Organ-specific aging clocks were derived in their second-generation form from Goeminne et al. (19) and were calculated following the authors’ instructions.

### Time-to-event analyses

We conducted three sets of time-to-event analyses, all based on Cox proportional hazards regression models as implemented in R using the *survival* package (version 3.8-3). Across these analyses, we assessed the association of proteomic aging clock estimates, used as predictors, with (i) all-cause mortality, (ii) mortality due to cancer (i.e. cancer as the underlying cause of death), and (iii) cancer incidence. For cancer mortality and incidence, Fine–Gray subdistribution hazard regression(22) was additionally performed to account for competing risks of non-cancer death and pre-diagnostic death, respectively.

Proteomic aging clock estimates were scaled to a mean of zero and variance of one in these analyses for ease of downstream comparisons. Follow-up began on the day each twin attended the clinic and provided blood samples and ended either at the occurrence of the event (death or cancer diagnosis) or on 31 December 2024, whichever came first. In analyses(ii) and (iii), participants with a cancer diagnosis prior to blood sampling were excluded as detailed above. Accordingly, these analyses assessed associations with either cancer-related mortality arising from cancers diagnosed after the start of follow-up or with incident cancer, rather than cancer prevalence. The mean follow-up of the twins was 10.6 years (SD ± 1.6). In the Cox regression models, sex and age at blood sampling (i.e. at the start of follow-up) were included as initial covariates. To account for potential confounding by known lifestyle risk factors, subsequent models were further adjusted for baseline body mass index (BMI)(23) as measured at the clinic visit, cigarette smoking status (current, former, never)(24), and alcohol consumption based on a quantity-frequency questionnaire (g/month)(25). Across all models, family relatedness was accounted for by specifying clustered effects using family identifiers, resulting in the calculation of robust standard errors.

Nominal p-values were corrected for multiple testing within each set of analyses using the Benjamini-Hochberg procedure. The proportional hazards assumption was systematically assessed using the *cox*.*zph()* function. Main text results are presented as hazard ratios with 95% confidence intervals, while complete model statistics, including regression coefficients, multiple testing correction-adjusted p-values and nominal p-values, are detailed in the Supplementary Material.

## Results

Baseline characteristics of the study population are summarized in Table 1. At the time of blood sampling, participants (N = 401, 59% female) had a mean age of 62.3 years (SD ± 3.8). Proteomic aging clock estimates showed varied correlations with chronological age, ranging from *R* = –0.38 (HPS; negative correlation is expected for healthspan) to *R* = 0.77 (ProtAge; Supplementary Figure S1). Cross-clock evaluations demonstrated strong inter-correlation among systemic clocks and identified a cluster of correlated organ-specific aging clocks, notably Brain, Immune, and Liver (Supplementary Figure S2).

**Table 1.**
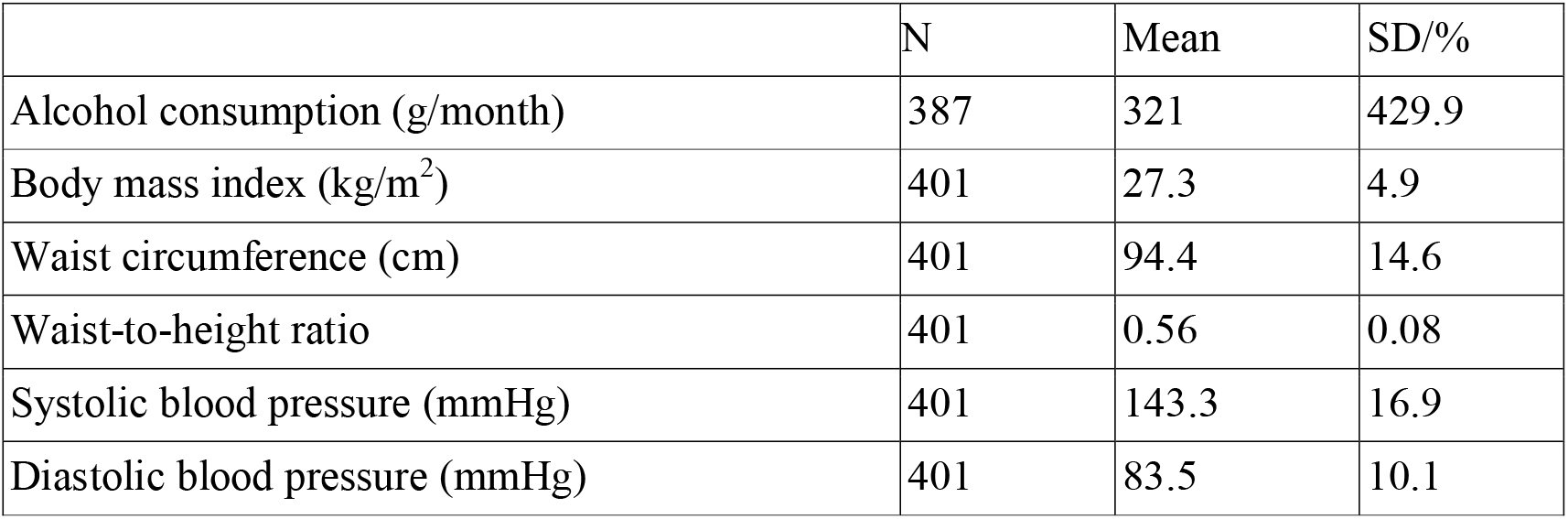
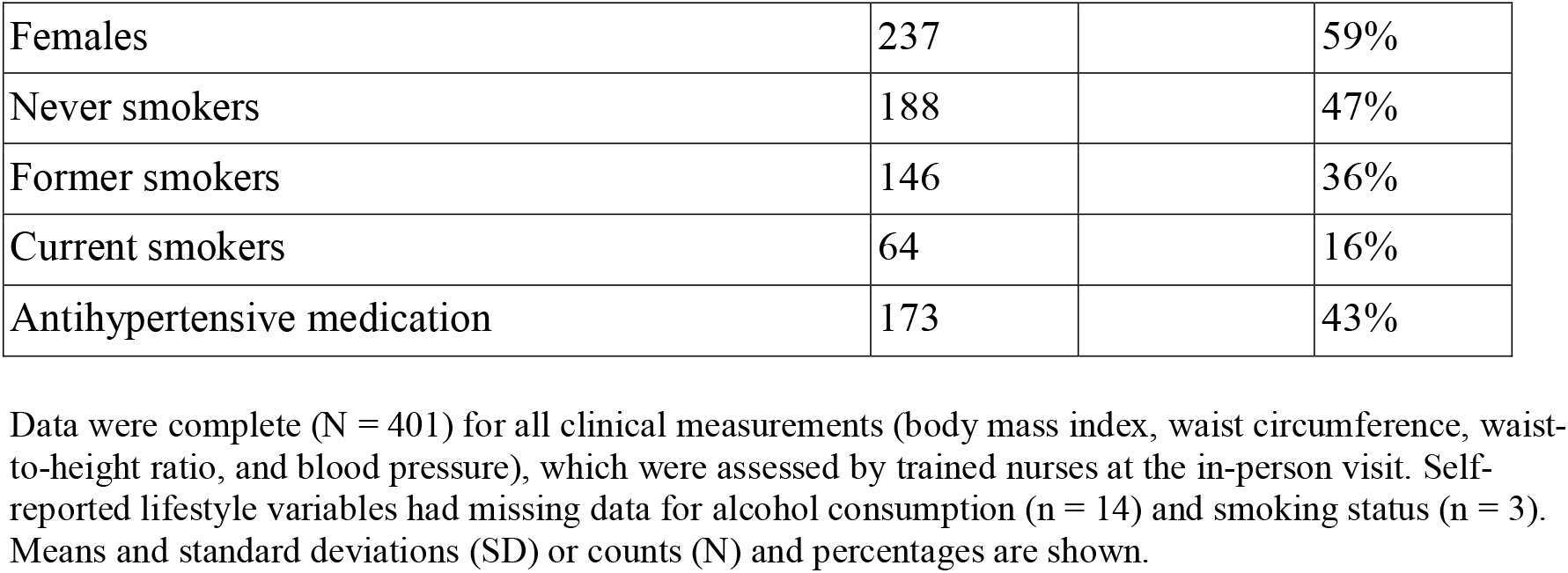
Descriptive statistics of the study sample at the start of the follow-up (i.e., at blood sampling).

|  | N | Mean | SD/% |
| --- | --- | --- | --- |
| Alcohol consumption (g/month) | 387 | 321 | 429.9 |
| Body mass index (kg/m<sup>2</sup>) | 401 | 27.3 | 4.9 |
| Waist circumference (cm) | 401 | 94.4 | 14.6 |
| Waist-to-height ratio | 401 | 0.56 | 0.08 |
| Systolic blood pressure (mmHg) | 401 | 143.3 | 16.9 |
| Diastolic blood pressure (mmHg) | 401 | 83.5 | 10.1 |
| Females | 237 |  | 59% |
| Never smokers | 188 |  | 47% |
| Former smokers | 146 |  | 36% |
| Current smokers | 64 |  | 16% |
| Antihypertensive medication | 173 |  | 43% |

Data were complete (N = 401) for all clinical measurements (body mass index, waist circumference, waist-to-height ratio, and blood pressure), which were assessed by trained nurses at the in-person visit. Self-reported lifestyle variables had missing data for alcohol consumption (n = 14) and smoking status (n = 3). Means and standard deviations (SD) or counts (N) and percentages are shown.

In the basic Cox proportional hazards regression models adjusted for age and sex only, estimates from 15 of the 22 proteomic aging clocks were significantly associated with all-cause mortality after multiple testing correction (FDR *p* < 0.05; Figure 1A, Table 2). Among systemic measures, higher Conventional (HR = 2.64, 95% CI: 1.89–3.69) and PAC (HR = 2.49, 95% CI: 1.79–3.47) scores were associated with an approximate 2.5-fold increase in mortality risk per 1 SD increment, whereas higher HPS showed a strong inverse association with mortality (HR = 0.50, 95% CI: 0.40–0.64), which was expected as higher values indicate higher expected healthspan. In line with this, Kaplan–Meier survival curves stratified by HPS tertiles showed that mortality was concentrated primarily in the lowest healthspan tertile (Figure 1B). Twelve organ-specific aging clocks were also significantly associated with all-cause mortality in basic models, with the strongest risks observed for the Immune (HR = 2.27, 95% CI: 1.80–2.87), Brain (HR = 2.04, 95% CI: 1.36–3.04), and Liver (HR = 2.00, 95% CI: 1.50–2.68) aging clocks. Subsequent adjustment for lifestyle risk factors (BMI, smoking status, and alcohol consumption) attenuated associations across the board, but 11 aging clocks remained predictive of mortality (FDR *p* < 0.05; Table 2). These included three systemic aging clocks (Conventional, PAC, and HPS) and eight organ-specific aging clocks (Immune, Liver, Artery, Intestine, Adipose, Kidney, Pancreas, and Skin). The associations for the Brain, Stomach, Esophagus, and Adrenal aging clocks were attenuated to non-significance.

**Table 2.**
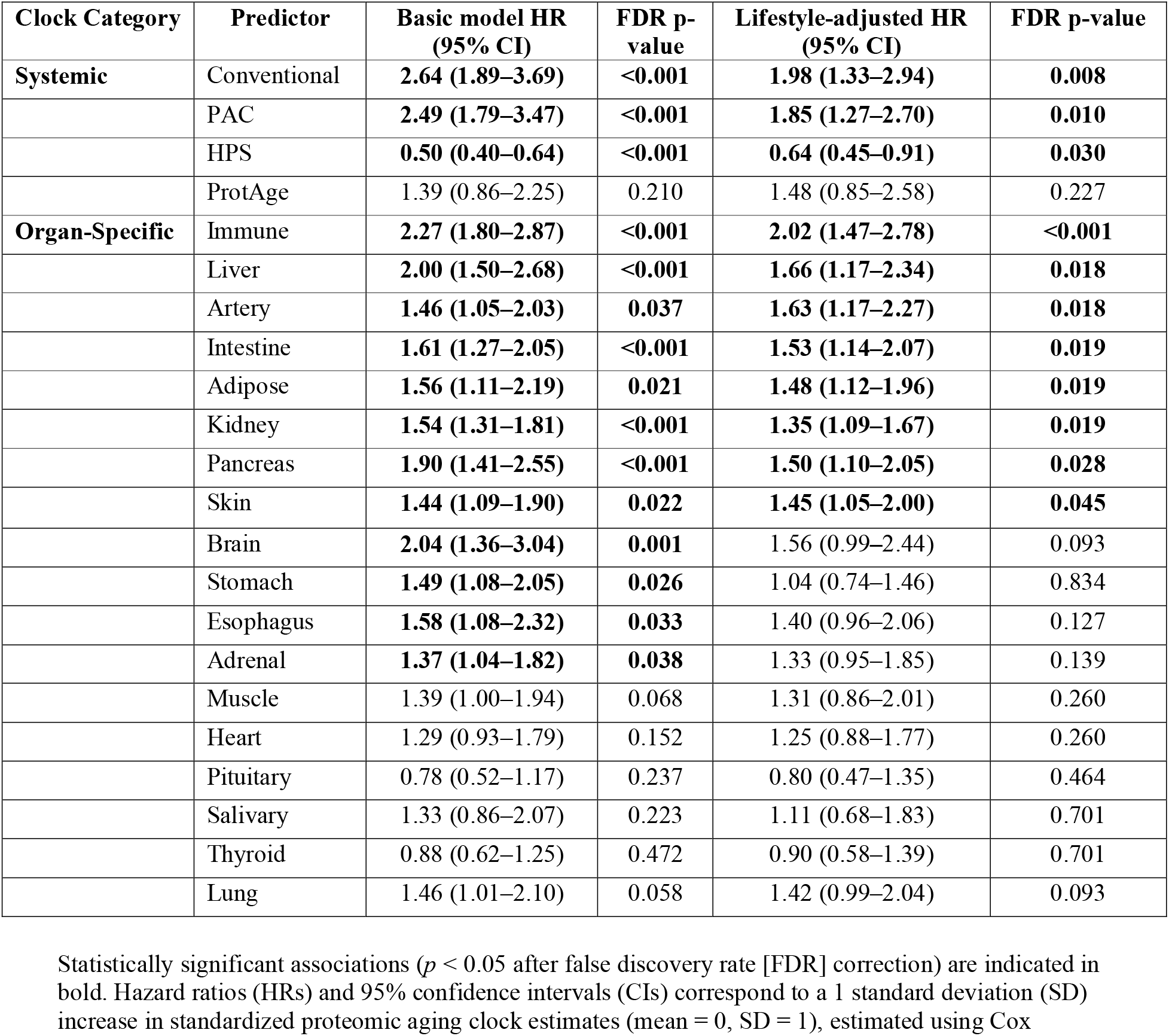
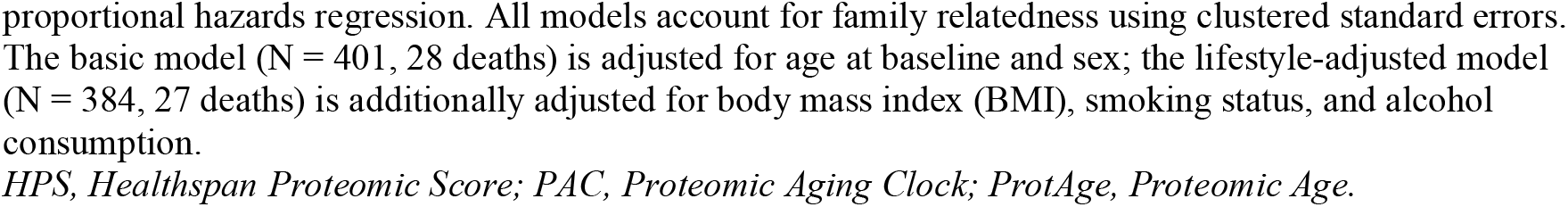
Associations between systemic and organ-specific proteomic aging clock estimates and all-cause mortality.

| Clock Category | Predictor | Basic model HR<br>(95% CI) | FDR p-value | Lifestyle-adjusted HR<br>(95% CI) | FDR p-value |
| --- | --- | --- | --- | --- | --- |
| **Systemic** | Conventional | **2.64 (1.89–3.69)** | **<0.001** | **1.98 (1.33–2.94)** | **0.008** |
|  | PAC | **2.49 (1.79–3.47)** | **<0.001** | **1.85 (1.27–2.70)** | **0.010** |
|  | HPS | **0.50 (0.40–0.64)** | **<0.001** | **0.64 (0.45–0.91)** | **0.030** |
|  | ProtAge | 1.39 (0.86–2.25) | 0.210 | 1.48 (0.85–2.58) | 0.227 |
| **Organ-Specific** | Immune | **2.27 (1.80–2.87)** | **<0.001** | **2.02 (1.47–2.78)** | **<0.001** |
|  | Liver | **2.00 (1.50–2.68)** | **<0.001** | **1.66 (1.17–2.34)** | **0.018** |
|  | Artery | **1.46 (1.05–2.03)** | **0.037** | **1.63 (1.17–2.27)** | **0.018** |
|  | Intestine | **1.61 (1.27–2.05)** | **<0.001** | **1.53 (1.14–2.07)** | **0.019** |
|  | Adipose | **1.56 (1.11–2.19)** | **0.021** | **1.48 (1.12–1.96)** | **0.019** |
|  | Kidney | **1.54 (1.31–1.81)** | **<0.001** | **1.35 (1.09–1.67)** | **0.019** |
|  | Pancreas | **1.90 (1.41–2.55)** | **<0.001** | **1.50 (1.10–2.05)** | **0.028** |
|  | Skin | **1.44 (1.09–1.90)** | **0.022** | **1.45 (1.05–2.00)** | **0.045** |
|  | Brain | **2.04 (1.36–3.04)** | **0.001** | 1.56 (0.99–2.44) | 0.093 |
|  | Stomach | **1.49 (1.08–2.05)** | **0.026** | 1.04 (0.74–1.46) | 0.834 |
|  | Esophagus | **1.58 (1.08–2.32)** | **0.033** | 1.40 (0.96–2.06) | 0.127 |
|  | Adrenal | **1.37 (1.04–1.82)** | **0.038** | 1.33 (0.95–1.85) | 0.139 |
|  | Muscle | 1.39 (1.00–1.94) | 0.068 | 1.31 (0.86–2.01) | 0.260 |
|  | Heart | 1.29 (0.93–1.79) | 0.152 | 1.25 (0.88–1.77) | 0.260 |
|  | Pituitary | 0.78 (0.52–1.17) | 0.237 | 0.80 (0.47–1.35) | 0.464 |
|  | Salivary | 1.33 (0.86–2.07) | 0.223 | 1.11 (0.68–1.83) | 0.701 |
|  | Thyroid | 0.88 (0.62–1.25) | 0.472 | 0.90 (0.58–1.39) | 0.701 |
|  | Lung | 1.46 (1.01–2.10) | 0.058 | 1.42 (0.99–2.04) | 0.093 |

Statistically significant associations ( $p < 0.05$ after false discovery rate [FDR] correction) are indicated in bold. Hazard ratios (HRs) and 95% confidence intervals (CIs) correspond to a 1 standard deviation (SD) increase in standardized proteomic aging clock estimates (mean = 0, SD = 1), estimated using Cox
proportional hazards regression. All models account for family relatedness using clustered standard errors. The basic model (N = 401, 28 deaths) is adjusted for age at baseline and sex; the lifestyle-adjusted model (N = 384, 27 deaths) is additionally adjusted for body mass index (BMI), smoking status, and alcohol consumption.
*HPS, Healthspan Proteomic Score; PAC, Proteomic Aging Clock; ProtAge, Proteomic Age.*

**Figure 1.**
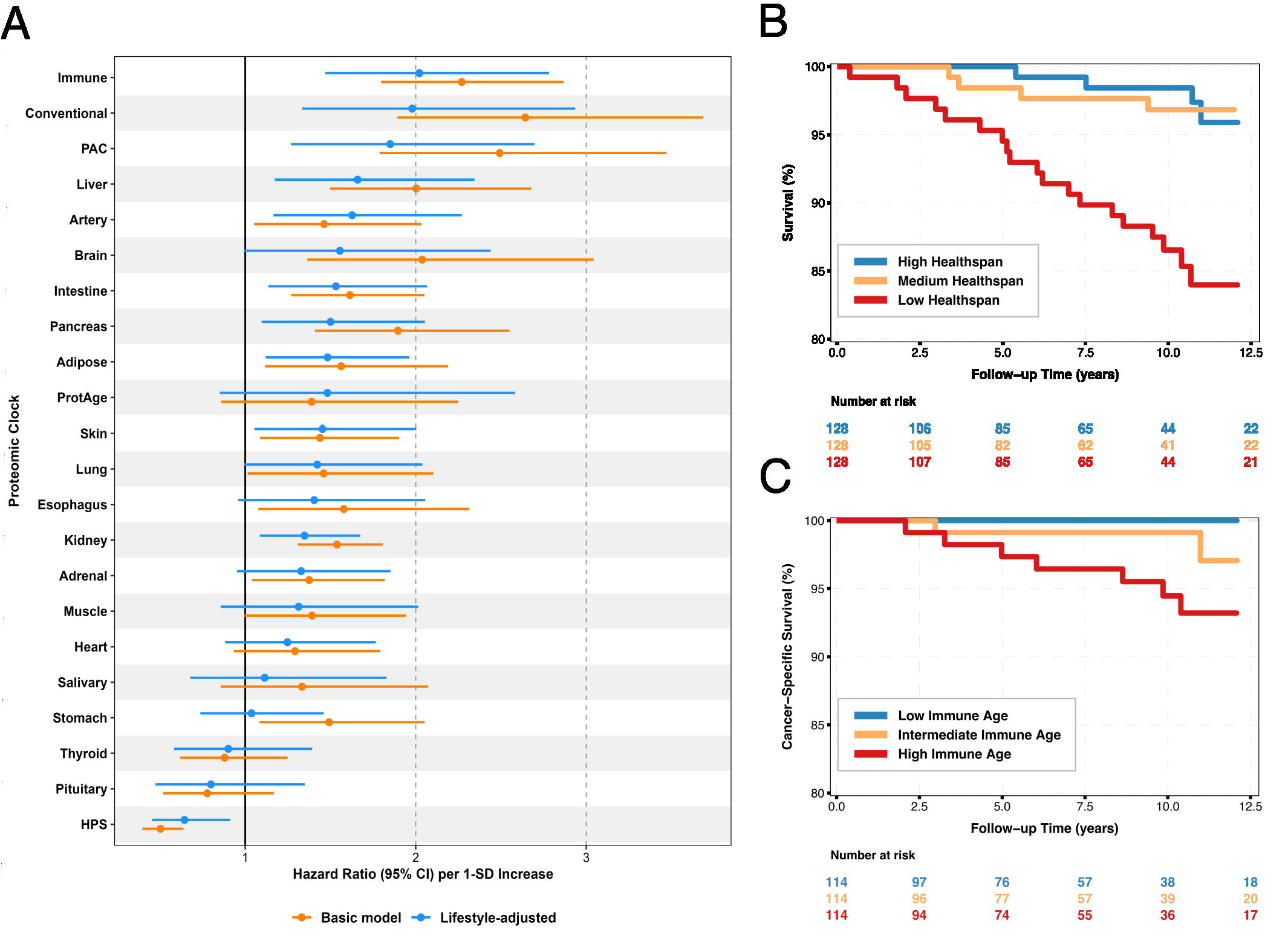
Systemic and organ-specific proteomic aging clocks in relation to survival outcomes. (A) Forest plot showing hazard ratios (HR) and 95% confidence intervals (CI) per one standard deviation (1-SD) increase across systemic and organ-specific proteomic aging clocks. Estimates are displayed for both the basic model (orange; adjusted for age and sex; N = 401, 28 deaths) and the lifestyle-adjusted model (blue; additionally adjusted for body mass index, smoking status, and alcohol consumption; N = 384, 27 deaths), with the vertical reference line at HR = 1.0 indicating no difference in risk. Kaplan–Meier survival curves over 12 years of follow-up stratified by Healthspan Proteomic Score (HPS) tertiles (High, Medium, and Low Healthspan; N = 384, n = 128 per tertile). The table below indicates the number of individuals at risk across 2.5-year intervals. Kaplan–Meier cancer-specific survival curves over 12 years of follow-up stratified by Immune Age (Low, Intermediate, and High Immune Age, N = 342, n = 114 per tertile, 9 deaths from cancer). The table below indicates the number of individuals at risk across 2.5-year intervals. *CI, confidence interval; HPS, Healthspan Proteomic Score; HR, hazard ratio; PAC, Proteomic Aging Clock; ProtAge, Proteomic Age; SD, standard deviation*.

When predicting cancer-specific mortality among cancer-free individuals at baseline (N = 355, 9 events), the Immune aging clock was the only predictor significantly associated with the outcome after multiple testing correction (basic model HR = 2.94, 95% CI: 1.88–4.59, DR *p* < 0.001). This association remained robust and strengthened after multivariable adjustment for lifestyle risk factors (HR = 3.31, 95% CI: 1.78–6.17, FDR *p* = 0.004).

Kaplan–Meier survival analyses supported these findings and showed a pronounced risk gradient across Immune clock tertiles, where subjects in the highest tertile accounted for most events, whereas no deaths from cancer occurred in the lowest tertile over follow-up (Figure 1C). Although several other systemic and organ-specific aging clocks displayed consistent directional trends, where higher proteomic age and lower healthspan seemed to be linked to elevated cancer mortality risk, none met the threshold for statistical significance following FDR adjustment (all FDR *p* > 0.05; Supplementary Table S2).

In analyses of cancer incidence (34 events), nominal associations (*p* < 0.05) were observed for five organ-specific aging clocks in basic models (Kidney, Muscle, Immune, Adrenal, Intestine) and persisted for four (excluding Intestine) after lifestyle adjustment (31 events), all reflecting a lower risk of incident cancer per standard deviation increase in proteomic age (Supplementary Table S3). However, none of these associations survived multiple testing correction (all FDR *p* > 0.05).

The proportional hazards assumption was satisfied across all models of all-cause and cancer-specific mortality and was largely met for cancer incidence (all residual tests detailed in Supplementary Tables S1–S3).

Furthermore, sensitivity analyses using Fine-Gray subdistribution hazard models to account for competing mortality yielded consistent findings for cancer mortality and confirmed null associations for cancer incidence (Supplementary Table S4).

## Discussion

In the present study, we evaluated the predictive performance of a panel of systemic and organ-specific proteomic aging clocks for all-cause mortality, cancer-related mortality, and cancer incidence using Finnish twin individuals. Our results indicate that proteomic aging clocks robustly predict all-cause mortality, even in samples of modest size such as the one we examined, with several aging clocks — notably the systemic Conventional and PAC clocks, as well as the Immune, Brain, and Liver organ-specific aging clocks — exhibiting a doubling of mortality risk per standard deviation increment in clock estimates. Although the performance of proteomic aging clocks in predicting cancer incidence was limited, proteomic aging clocks were more informative for predicting cancer-related mortality, even with a relatively small number of cancer-related deaths occurring after the start of follow-up.

Notably, the Immune clock exhibited particularly strong predictive performance for cancer-related mortality. This finding aligns with a growing body of evidence demonstrating that chronic immune-system aging, often termed ‘inflammaging’(26), and T-cell senescence actively drive tumor progression and impair clinical outcomes(27). Taken together, these findings suggest that proteomic aging clocks, particularly the Immune clock, may be useful for identifying individuals at elevated risk of developing cancers associated with high mortality.

Interestingly, while the proteomic aging clocks robustly predicted all-cause mortality and the Immune clock strongly predicted cancer mortality, they largely failed to predict incident cancer, with several clocks even displaying nominal inverse associations. Similar findings have been recently documented in larger cohorts. For instance, a study of over 22,000 participants in an Italian population cohort found that while epigenetic age acceleration strongly predicted all-cause mortality, it was paradoxically associated with a reduced risk of incident breast and prostate cancers(28). Similar inverse associations between organ-specific proteomic aging clocks and incident cancers were noted in the Whitehall II cohort(29).

Conversely, although recent work by Tian et al. did identify positive associations between biological aging—including the Immune clock—and incident cancer, these associations were largely confined to cancers manifesting earlier in life (<55 years) and attenuated with older age(30). Importantly, while Fine–Gray sensitivity analyses indicated that these nominal inverse associations largely attenuated once accounting for competing mortality, they confirmed a striking biological decoupling between cancer initiation and lethality. This may be explained by the antagonistic relationship between aging and oncogenesis. Mechanisms such as cellular senescence, which contribute to biological age acceleration and organ dysfunction, simultaneously serve as potent tumor-suppressive barriers by permanently arresting the proliferation of damaged cells(31). Metabolic silencing may also play a role, where “old” proteomic profiles of organs often reflect metabolic exhaustion or decreased growth factor signaling, which are hostile to the rapid energy demands of an emerging tumor(32).

Despite the strengths of our study, including the comprehensive profiling of 22 distinct proteomic aging clocks, complete registry-based follow-up without loss to attrition, and methodological control for lifestyle confounders, limitations should be acknowledged. The most notable limitation is the relatively modest sample size, which resulted in a low number of deaths and cancer diagnoses over the follow-up period. This likely limited our ability to detect additional associations between proteomic aging clocks and survival or cancer incidence, with many nominal associations failing to pass correction for multiple testing. This also prevented us from stratifying analyses per cancer type, and our findings do not incorporate a large diversity of cancer types either given the limited number of deaths and cancer onset observed. Additionally, we could not conduct any pairwise analyses due to the small number of cancer cases. Cancer incidence and mortality in the Finnish Twin Cohort do not differ from the population at large, indicating that our results are likely generalizable(33).

Furthermore, the mean age at the end of follow-up was just over 70 years, which does not capture very old age in the Finnish population. Consequently, many individuals who may ultimately be diagnosed with cancer or die from cancer were still alive at the end of follow-up. A longer follow-up period would not only increase statistical power by capturing more cancer-related deaths but also allow inclusion of cancers that manifest primarily in very old age. Further studies are needed both to replicate our findings in larger cohorts and to elucidate the biological mechanisms underlying the observed associations.

In conclusion, our study indicates that proteomic aging clocks are good predictors of all-cause and cancer-related deaths, even in samples of modest size. Our study thus suggests proteomic aging clocks may be useful for public health purposes, notably for early prevention of high-mortality cancers, with further studies needed to fully uncover the potential of these clocks, especially through stratification by cancer types.

## Data Availability

The Finnish Twin Cohort data used in the analysis are deposited in the Biobank of the Finnish Institute for Health and Welfare. Access to Finnish Cancer Registry data can be applied for as part of the Biobank access procedures. These data are available for use by qualified researchers through a standardized application procedure (https://findata.fi/en/permits/). Further information is available from the corresponding author upon request.

## Acknowledgments and further disclosures

We want to acknowledge the participants and investigators of the FinnGen study, which is a genomics and personalized medicine research project that provided data resources, particularly Olink proteomics in twins of EH-Epi, for the completion of this study.

## Ethics statement

The study protocol was approved by the Institutional Ethics Board of the Hospital District of Helsinki and Uusimaa, Finland (ID 154/13/03/00/11). Permission for linkage to the Finnish Cancer Registry was provided by the Finnish Social and Health Data Permit Authority Findata (THL/6353/14.06.00/2023). Blood samples were collected from each participant after they had signed a written informed consent.

## Authors contributions

All authors contributed to the conceptualization and methodology of the present study. XW contributed to resources and data curation. JK and MO acquired the data. VL and GD conducted the statistical analyses, interpreted the results and wrote the original draft; all authors contributed to interpreting the findings and critically reviewing and editing the manuscript. GD, MO and JK supervised the study. VL finalized the manuscript, and all authors read and approved the final manuscript.

## Abbreviations

BMI: Body mass index
CI: Confidence interval
C-index: Harrell’s concordance index
DNAm: DNA methylation
EH-Epi: Essential Hypertension Epigenetic study
FDR: False discovery rate
FTC: Finnish Twin Cohort
HPS: Healthspan Proteomic Score
HR: Hazard Ratio
PAC: Proteomic Aging Clock
ProtAge: Proteomic Age
SD: Standard deviation
SHR: Subdistribution hazard ratio
UKB-PPP: UK Biobank Pharma Proteomics Project

