## Supplementary material for "Systemic and organ-specific proteomic aging clocks as predictors of cancer incidence and survival: insights from the Finnish Twin Cohort"

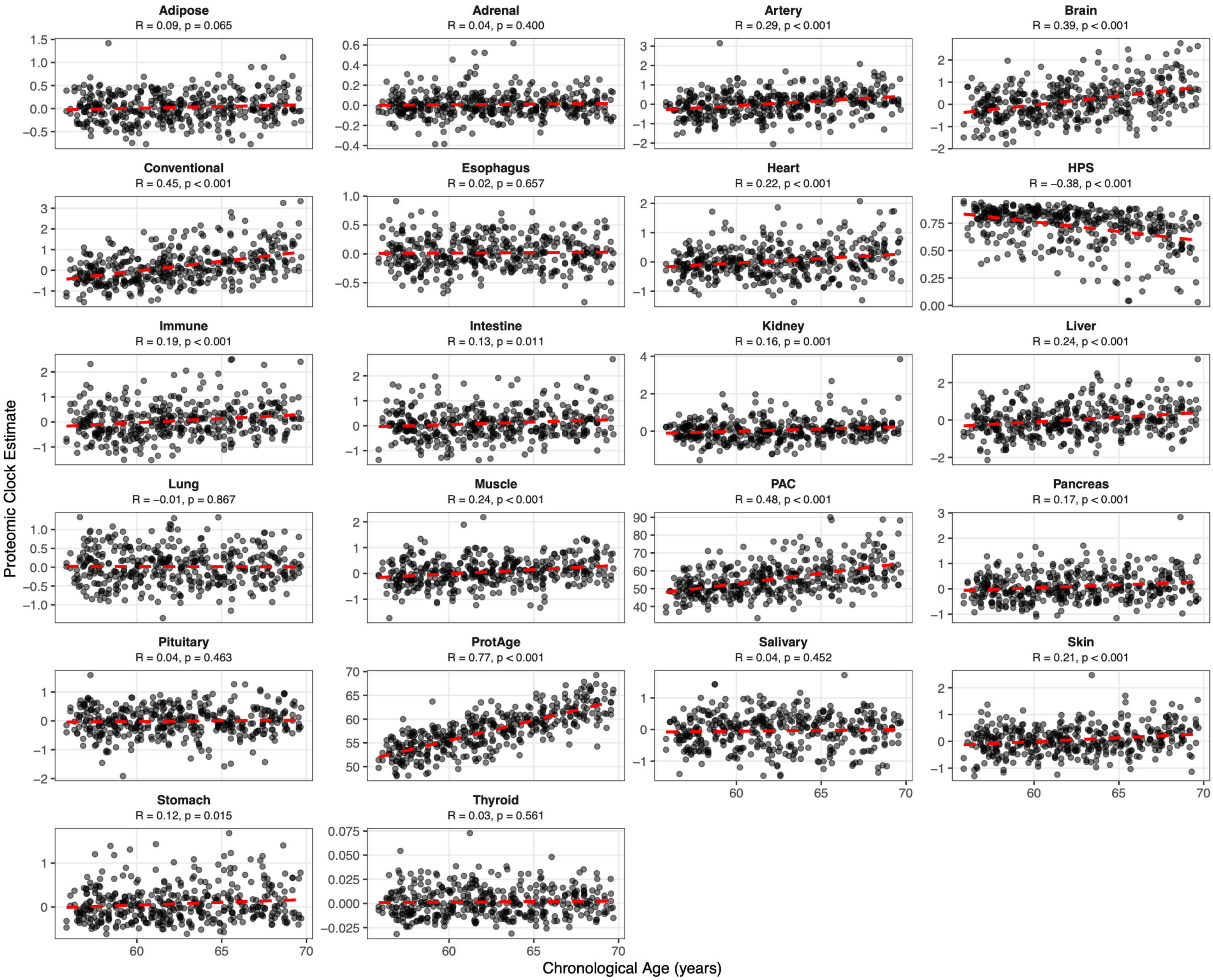


**Figure S1.** Associations between systemic and organ-specific proteomic clock estimates and chronological age.

Scatter plots displaying baseline chronological age (x-axis, years) versus 22 systemic and organ-specific proteomic clock estimates (y-axis). Solid red lines represent linear regression fits. Pearson correlation coefficients (R) and corresponding *p*-values are reported for each panel. Y-axis scales vary across panels to accommodate the native score metrics of each respective clock algorithm (e.g., predicted age in years for ProtAge and PAC; model-derived index scores for organ-specific clocks; composite score for HPS).
*HPS, Healthspan Proteomic Score; PAC, Proteomic Age Clock; R, Pearson correlation coefficient.*

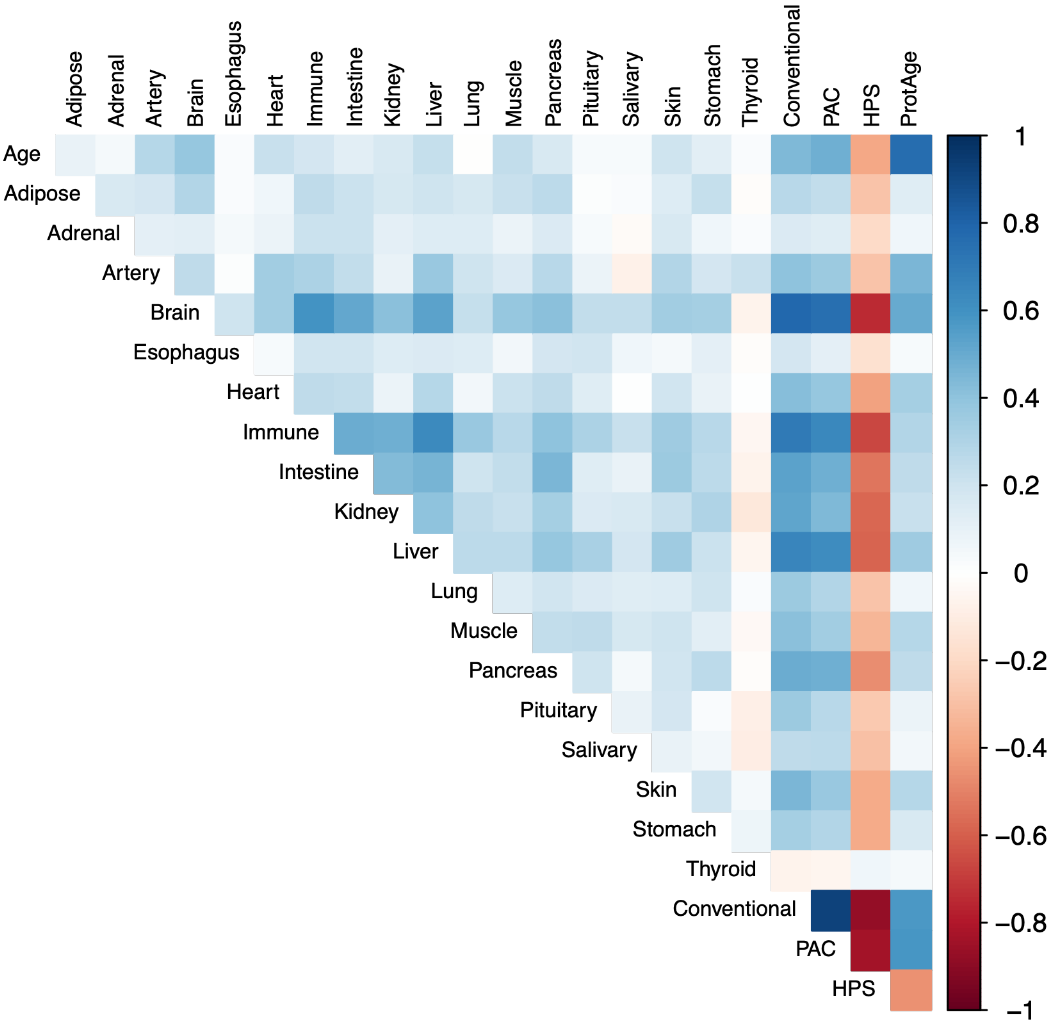


**Figure S2.** Pairwise correlation matrix of systemic and organ-specific proteomic clocks and chronological age.

Heatmap displaying pairwise Pearson correlation coefficients between baseline chronological age and 22 systemic and organ-specific proteomic clock estimates. Tile colors represent the magnitude and direction of the correlation according to the color gradient bar: shades of blue indicate positive correlations, shades of red indicate inverse correlations, and white indicates near-zero correlation. Systemic clocks trained on chronological age (ProtAge) or mortality (Conventional, PAC) correlate positively with age, whereas the Healthspan Proteomic Score (HPS) reflects healthspan and displays inverse correlations with chronological age and the other proteomic clocks.
*HPS, Healthspan Proteomic Score; PAC, Proteomic Age Clock.*

**Table S1.** Associations between systemic and organ-specific proteomic clocks and all-cause mortality: Cox proportional hazards regression models and diagnostics.

| | | Basic Model (N = 401, Events = 28) | | | | | | Lifestyle-Adjusted Model (N = 384, Events = 27) | | | | | |
|---|---|---|---|---|---|---|---|---|---|---|---|---|---|
| **Clock Category** | **Predictor** | **Beta** | **HR (95% CI)** | **p-value** | **FDR p-value** | **ZPH** | **C-index** | **Beta** | **HR (95% CI)** | **p-value** | **FDR p-value** | **ZPH** | **C-index** |
| **Systemic** | Conventional | 0.97 | **2.64 (1.89–3.69)** | **<0.001** | **<0.001** | 0.95 | 0.83 | 0.68 | **1.98 (1.33–2.94)** | **<0.001** | **0.008** | 0.79 | 0.84 |
| | PAC | 0.91 | **2.49 (1.79–3.47)** | **<0.001** | **<0.001** | 0.87 | 0.82 | 0.62 | **1.85 (1.27–2.70)** | **0.001** | **0.010** | 0.81 | 0.84 |
| | HPS | -0.69 | **0.50 (0.40–0.64)** | **<0.001** | **<0.001** | 0.86 | 0.82 | -0.44 | **0.64 (0.45–0.91)** | **0.013** | **0.030** | 0.77 | 0.84 |
| | ProtAge | 0.33 | 1.39 (0.86–2.25) | 0.181 | 0.210 | 0.18 | 0.77 | 0.39 | 1.48 (0.85–2.58) | 0.165 | 0.227 | 0.49 | 0.84 |
| **Organ-Specific** | Immune | 0.82 | **2.27 (1.80–2.87)** | **<0.001** | **<0.001** | 0.51 | 0.82 | 0.70 | **2.02 (1.47–2.78)** | **<0.001** | **<0.001** | 0.48 | 0.86 |
| | Liver | 0.69 | **2.00 (1.50–2.68)** | **<0.001** | **<0.001** | 0.90 | 0.81 | 0.51 | **1.66 (1.17–2.34)** | **0.004** | **0.018** | 0.75 | 0.84 |
| | Artery | 0.38 | **1.46 (1.05–2.03)** | **0.024** | **0.037** | 0.78 | 0.80 | 0.49 | **1.63 (1.17–2.27)** | **0.004** | **0.018** | 0.67 | 0.85 |
| | Intestine | 0.48 | **1.61 (1.27–2.05)** | **<0.001** | **<0.001** | 0.68 | 0.79 | 0.43 | **1.53 (1.14–2.07)** | **0.005** | **0.019** | 0.60 | 0.84 |
| | Adipose | 0.45 | **1.56 (1.11–2.19)** | **0.010** | **0.021** | 0.18 | 0.79 | 0.39 | **1.48 (1.12–1.96)** | **0.006** | **0.019** | 0.56 | 0.84 |
| | Kidney | 0.43 | **1.54 (1.31–1.81)** | **<0.001** | **<0.001** | 0.07 | 0.80 | 0.30 | **1.35 (1.09–1.67)** | **0.007** | **0.019** | 0.25 | 0.85 |
| | Pancreas | 0.64 | **1.90 (1.41–2.55)** | **<0.001** | **<0.001** | 0.69 | 0.80 | 0.41 | **1.50 (1.10–2.05)** | **0.011** | **0.028** | 0.60 | 0.84 |
| | Skin | 0.36 | **1.44 (1.09–1.90)** | **0.011** | **0.022** | 0.68 | 0.79 | 0.37 | **1.45 (1.05–2.00)** | **0.023** | **0.045** | 0.70 | 0.84 |
| | Brain | 0.71 | **2.04 (1.36–3.04)** | **<0.001** | **0.001** | 0.87 | 0.80 | 0.44 | 1.56 (0.99–2.44) | 0.055 | 0.093 | 0.78 | 0.84 |
| | Stomach | 0.40 | **1.49 (1.08–2.05)** | **0.014** | **0.026** | 0.72 | 0.78 | 0.04 | 1.04 (0.74–1.46) | 0.834 | 0.834 | 0.70 | 0.84 |
| | Esophagus | 0.46 | **1.58 (1.08–2.31)** | **0.019** | **0.033** | 0.81 | 0.79 | 0.34 | 1.40 (0.96–2.06) | 0.081 | 0.127 | 0.67 | 0.85 |
| | Adrenal | 0.32 | **1.37 (1.04–1.82)** | **0.026** | **0.038** | 0.83 | 0.77 | 0.28 | 1.33 (0.95–1.85) | 0.095 | 0.139 | 0.73 | 0.84 |
| | Muscle | 0.33 | 1.39 (1.00–1.94) | 0.052 | 0.068 | 0.69 | 0.78 | 0.27 | 1.31 (0.86–2.01) | 0.213 | 0.260 | 0.66 | 0.84 |
| | Heart | 0.26 | 1.29 (0.93–1.79) | 0.124 | 0.152 | 0.86 | 0.78 | 0.22 | 1.25 (0.88–1.77) | 0.211 | 0.260 | 0.77 | 0.85 |
| | Pituitary | -0.25 | 0.78 (0.52–1.17) | 0.226 | 0.237 | 0.60 | 0.77 | -0.22 | 0.80 (0.47–1.35) | 0.401 | 0.464 | 0.48 | 0.84 |
| | Salivary | 0.29 | 1.33 (0.86–2.07) | 0.203 | 0.223 | 0.67 | 0.78 | 0.11 | 1.11 (0.68–1.83) | 0.669 | 0.701 | 0.67 | 0.84 |
| | Thyroid | -0.13 | 0.88 (0.62–1.25) | 0.472 | 0.472 | 0.80 | 0.77 | -0.10 | 0.90 (0.58–1.39) | 0.638 | 0.701 | 0.65 | 0.84 |
| | Lung | 0.38 | 1.46 (1.01–2.10) | 0.042 | 0.058 | 0.80 | 0.78 | 0.35 | 1.42 (0.99–2.04) | 0.055 | 0.093 | 0.73 | 0.85 |

Hazard ratios (HRs) and 95% confidence intervals (CIs) correspond to a 1-SD increase in each standardized proteomic clock estimate. Basic model adjusted for age and sex (N = 401, 28 deaths). Lifestyle-adjusted model additionally adjusted for body mass index (BMI), smoking status, and alcohol consumption (N = 384, 27 deaths; sample size reduction reflects missing covariate data). Bold values indicate statistically significant associations following correction for multiple testing across all clocks using the Benjamini–Hochberg false discovery rate (FDR $p < 0.05$). Proportional hazards assumptions were evaluated using Schoenfeld residuals (ZPH), where $p > 0.05$ indicates no violation of the assumption. Model discrimination was assessed using Harrell's concordance index (C-index).

*Beta, regression coefficient; CI, confidence interval; C-index, Harrell's concordance index; FDR, false discovery rate; HPS, Healthspan Proteomic Score; HR, hazard ratio; PAC, Proteomic Age Clock; SD, standard deviation; ZPH, test of proportional hazards based on Schoenfeld residuals.*

**Table S2.** Associations between systemic and organ-specific proteomic clocks and cancer-related mortality: Cox proportional hazards regression models and diagnostics.

| | | Basic Model (N = 355, Events = 9) | | | | | | Lifestyle-Adjusted (N = 342, Events = 9) | | | | | |
|---|---|---|---|---|---|---|---|---|---|---|---|---|---|
| **Clock Category** | **Predictor** | **Beta** | **HR (95% CI)** | **p-value** | **FDR p-value** | **ZPH** | **C-index** | **Beta** | **HR (95% CI)** | **p-value** | **FDR p-value** | **ZPH** | **C-index** |
| **Systemic** | Conventional | 0.66 | 1.93 (1.03–3.60) | 0.040 | 0.135 | 0.45 | 0.84 | 0.42 | 1.53 (0.87–2.68) | 0.138 | 0.339 | 0.49 | 0.88 |
| | PAC | 0.61 | 1.84 (0.88–3.85) | 0.104 | 0.190 | 0.41 | 0.82 | 0.40 | 1.49 (0.79–2.82) | 0.222 | 0.376 | 0.50 | 0.87 |
| | HPS | -0.51 | 0.60 (0.38–0.95) | 0.028 | 0.122 | 0.42 | 0.84 | -0.37 | 0.69 (0.45–1.06) | 0.093 | 0.339 | 0.50 | 0.88 |
| | ProtAge | 0.50 | 1.65 (0.75–3.64) | 0.217 | 0.318 | 0.35 | 0.81 | 0.43 | 1.54 (0.60–3.95) | 0.372 | 0.518 | 0.55 | 0.87 |
| **Organ-Specific** | Immune | 1.08 | **2.94 (1.88–4.59)** | **<0.001** | **<0.001** | 0.32 | 0.89 | 1.20 | **3.31 (1.78–6.17)** | **<0.001** | **0.004** | 0.49 | 0.91 |
| | Liver | 0.67 | 1.95 (0.98–3.87) | 0.057 | 0.156 | 0.39 | 0.84 | 0.61 | 1.85 (0.84–4.06) | 0.127 | 0.339 | 0.54 | 0.88 |
| | Artery | 0.25 | 1.29 (0.78–2.14) | 0.327 | 0.379 | 0.35 | 0.81 | 0.22 | 1.25 (0.72–2.16) | 0.429 | 0.525 | 0.58 | 0.87 |
| | Intestine | 0.28 | 1.33 (0.88–2.00) | 0.182 | 0.285 | 0.40 | 0.79 | 0.32 | 1.37 (0.86–2.19) | 0.185 | 0.339 | 0.59 | 0.87 |
| | Adipose | 0.53 | 1.71 (1.07–2.72) | 0.025 | 0.122 | 0.25 | 0.84 | 0.33 | 1.39 (0.89–2.16) | 0.147 | 0.339 | 0.48 | 0.89 |
| | Kidney | 0.32 | 1.38 (0.90–2.10) | 0.140 | 0.237 | 0.14 | 0.82 | 0.36 | 1.44 (0.95–2.19) | 0.088 | 0.339 | 0.38 | 0.88 |
| | Pancreas | 0.67 | 1.95 (1.10–3.47) | 0.023 | 0.122 | 0.31 | 0.86 | 0.48 | 1.62 (1.03–2.54) | 0.037 | 0.339 | 0.46 | 0.89 |
| | Skin | 0.30 | 1.36 (1.01–1.82) | 0.043 | 0.135 | 0.29 | 0.81 | 0.24 | 1.27 (0.90–1.80) | 0.180 | 0.339 | 0.50 | 0.88 |
| | Brain | 0.64 | 1.89 (0.88–4.03) | 0.100 | 0.190 | 0.36 | 0.82 | 0.38 | 1.46 (0.72–2.95) | 0.291 | 0.458 | 0.57 | 0.88 |
| | Stomach | 0.30 | 1.35 (0.81–2.27) | 0.253 | 0.327 | 0.18 | 0.81 | 0.12 | 1.13 (0.65–1.97) | 0.673 | 0.779 | 0.47 | 0.88 |
| | Esophagus | 0.42 | 1.52 (0.74–3.10) | 0.250 | 0.327 | 0.30 | 0.80 | 0.32 | 1.37 (0.68–2.78) | 0.377 | 0.518 | 0.55 | 0.88 |
| | Adrenal | 0.42 | 1.52 (0.96–2.39) | 0.072 | 0.157 | 0.34 | 0.81 | 0.41 | 1.50 (0.96–2.36) | 0.075 | 0.339 | 0.57 | 0.87 |
| | Muscle | 0.46 | 1.58 (0.97–2.59) | 0.069 | 0.157 | 0.35 | 0.80 | 0.40 | 1.49 (0.84–2.64) | 0.168 | 0.339 | 0.54 | 0.88 |
| | Heart | 0.10 | 1.11 (0.65–1.89) | 0.710 | 0.744 | 0.28 | 0.80 | 0.06 | 1.07 (0.52–2.17) | 0.859 | 0.868 | 0.50 | 0.87 |
| | Pituitary | -0.28 | 0.75 (0.43–1.32) | 0.323 | 0.379 | 0.33 | 0.79 | -0.31 | 0.74 (0.35–1.55) | 0.421 | 0.525 | 0.52 | 0.87 |
| | Salivary | 0.23 | 1.26 (0.67–2.36) | 0.467 | 0.513 | 0.37 | 0.80 | -0.07 | 0.94 (0.52–1.67) | 0.822 | 0.868 | 0.55 | 0.88 |
| | Thyroid | 0.02 | 1.02 (0.52–2.02) | 0.954 | 0.954 | 0.17 | 0.80 | -0.06 | 0.94 (0.48–1.85) | 0.868 | 0.868 | 0.40 | 0.88 |
| | Lung | 0.77 | 2.17 (1.16–4.05) | 0.015 | 0.122 | 0.36 | 0.87 | 0.63 | 1.89 (0.99–3.60) | 0.054 | 0.339 | 0.57 | 0.90 |

Hazard ratios (HRs) and 95% confidence intervals (CIs) correspond to a 1-SD increase in each standardized proteomic clock estimate. Basic model adjusted for age and sex (N = 355, 9 deaths from cancer). Lifestyle-adjusted model additionally adjusted for body mass index (BMI), smoking status, and alcohol consumption (N = 342, 9 deaths from cancer; sample size reduction reflects missing covariate data). Bold values indicate statistically significant associations following correction for multiple testing across all clocks using the Benjamini–Hochberg false discovery rate (FDR $p < 0.05$). Proportional hazards assumptions were evaluated using Schoenfeld residuals (ZPH), where $p > 0.05$ indicates no violation of the assumption. Model discrimination was assessed using Harrell's concordance index (C-index).
*Beta, regression coefficient; CI, confidence interval; C-index, Harrell's concordance index; FDR, false discovery rate; HPS, Healthspan Proteomic Score; HR, hazard ratio; PAC, Proteomic Age Clock; SD, standard deviation; ZPH, test of proportional hazards based on Schoenfeld residuals.*

**Table S3.** Associations between systemic and organ-specific proteomic clocks and cancer-incidence: Cox proportional hazards regression models and diagnostics.

| | | Basic Model (N = 355, Events = 34) | | | | | | Lifestyle-Adjusted Model (N = 342, Events = 31) | | | | | |
|---|---|---|---|---|---|---|---|---|---|---|---|---|---|
| **Clock Category** | **Predictor** | **Beta** | **HR (95% CI)** | **p-value** | **FDR p-value** | **ZPH** | **C-index** | **Beta** | **HR (95% CI)** | **p-value** | **FDR p-value** | **ZPH** | **C-index** |
| **Systemic** | Conventional | -0.17 | 0.85 (0.57–1.25) | 0.399 | 0.550 | 0.17 | 0.65 | -0.14 | 0.87 (0.49–1.55) | 0.642 | 0.784 | 0.18 | 0.64 |
| | PAC | -0.14 | 0.87 (0.60–1.28) | 0.485 | 0.617 | 0.12 | 0.66 | 0.00 | 1.00 (0.58–1.72) | 0.998 | 0.998 | 0.14 | 0.62 |
| | HPS | 0.11 | 1.11 (0.81–1.52) | 0.505 | 0.617 | 0.13 | 0.64 | 0.04 | 1.04 (0.46–2.34) | 0.921 | 0.964 | 0.12 | 0.62 |
| | ProtAge | -0.25 | 0.78 (0.49–1.23) | 0.281 | 0.476 | 0.10 | 0.65 | -0.22 | 0.80 (0.54–1.20) | 0.282 | 0.563 | 0.16 | 0.65 |
| **Organ-Specific** | Immune | -0.51 | 0.60 (0.40–0.90) | 0.013 | 0.070 | 0.09 | 0.67 | -0.83 | 0.44 (0.20–0.97) | 0.042 | 0.229 | 0.01 | 0.65 |
| | Liver | -0.03 | 0.97 (0.69–1.35) | 0.848 | 0.853 | 0.18 | 0.63 | 0.11 | 1.11 (0.76–1.63) | 0.590 | 0.784 | 0.16 | 0.63 |
| | Artery | -0.26 | 0.77 (0.51–1.18) | 0.234 | 0.469 | 0.09 | 0.63 | -0.37 | 0.69 (0.36–1.34) | 0.276 | 0.563 | 0.04 | 0.64 |
| | Intestine | -0.26 | 0.77 (0.60–0.99) | 0.039 | 0.171 | 0.29 | 0.64 | -0.40 | 0.67 (0.43–1.06) | 0.088 | 0.321 | 0.21 | 0.65 |
| | Adipose | -0.22 | 0.80 (0.50–1.28) | 0.350 | 0.550 | 0.14 | 0.64 | -0.29 | 0.75 (0.37–1.53) | 0.432 | 0.679 | 0.11 | 0.62 |
| | Kidney | -0.44 | 0.64 (0.48–0.85) | 0.002 | 0.051 | 0.10 | 0.68 | -0.65 | 0.52 (0.33–0.83) | 0.007 | 0.143 | 0.05 | 0.69 |
| | Pancreas | -0.36 | 0.70 (0.47–1.02) | 0.065 | 0.203 | 0.16 | 0.63 | -0.32 | 0.73 (0.35–1.51) | 0.392 | 0.664 | 0.07 | 0.62 |
| | Skin | 0.30 | 1.35 (1.00–1.82) | 0.051 | 0.187 | 0.19 | 0.63 | 0.22 | 1.25 (0.95–1.64) | 0.110 | 0.326 | 0.13 | 0.66 |
| | Brain | -0.29 | 0.75 (0.49–1.15) | 0.186 | 0.410 | 0.13 | 0.64 | -0.31 | 0.73 (0.39–1.37) | 0.328 | 0.601 | 0.05 | 0.66 |
| | Stomach | -0.45 | 0.64 (0.34–1.21) | 0.170 | 0.410 | 0.23 | 0.64 | -0.51 | 0.60 (0.34–1.06) | 0.079 | 0.321 | 0.12 | 0.67 |
| | Esophagus | 0.07 | 1.07 (0.80–1.45) | 0.643 | 0.745 | 0.21 | 0.64 | -0.02 | 0.98 (0.71–1.36) | 0.916 | 0.964 | 0.06 | 0.62 |
| | Adrenal | -0.30 | 0.74 (0.58–0.94) | 0.013 | 0.070 | 0.14 | 0.64 | -0.34 | 0.71 (0.55–0.93) | 0.014 | 0.149 | 0.31 | 0.63 |
| | Muscle | -0.42 | 0.66 (0.49–0.88) | 0.005 | 0.055 | 0.14 | 0.67 | -0.40 | 0.67 (0.47–0.95) | 0.023 | 0.166 | 0.19 | 0.67 |
| | Heart | -0.09 | 0.92 (0.53–1.57) | 0.753 | 0.829 | 0.11 | 0.63 | -0.07 | 0.94 (0.58–1.51) | 0.785 | 0.909 | 0.10 | 0.64 |
| | Pituitary | -0.17 | 0.84 (0.69–1.03) | 0.095 | 0.262 | 0.14 | 0.63 | -0.15 | 0.86 (0.71–1.04) | 0.124 | 0.326 | 0.16 | 0.65 |
| | Salivary | -0.05 | 0.95 (0.58–1.56) | 0.853 | 0.853 | 0.02 | 0.63 | 0.19 | 1.21 (0.54–2.70) | 0.637 | 0.784 | 0.00 | 0.62 |
| | Thyroid | 0.13 | 1.14 (0.84–1.54) | 0.400 | 0.550 | 0.18 | 0.62 | 0.41 | 1.51 (0.88–2.57) | 0.134 | 0.326 | 0.07 | 0.66 |
| | Lung | -0.30 | 0.74 (0.44–1.25) | 0.264 | 0.476 | 0.17 | 0.61 | -0.20 | 0.82 (0.44–1.52) | 0.520 | 0.763 | 0.19 | 0.64 |

Hazard ratios (HRs) and 95% confidence intervals (CIs) correspond to a 1-SD increase in each standardized proteomic clock estimate. Basic model adjusted for age and sex (N = 355, 34 incident cancer cases). Lifestyle-adjusted model additionally adjusted for body mass index (BMI), smoking status, and alcohol consumption (N = 342, 31 incident cancer cases; sample size reduction reflects missing covariate data). Bold values indicate statistically significant associations following correction for multiple testing across all clocks using the Benjamini–Hochberg false discovery rate (FDR $p < 0.05$). Proportional hazards assumptions were evaluated using Schoenfeld residuals (ZPH), where $p > 0.05$ indicates no violation of the assumption. Model discrimination was assessed using Harrell's concordance index (C-index).

*Beta, regression coefficient; CI, confidence interval; C-index, Harrell's concordance index; FDR, false discovery rate; HPS, Healthspan Proteomic Score; HR, hazard ratio; PAC, Proteomic Age Clock; SD, standard deviation; ZPH, test of proportional hazards based on Schoenfeld residuals.*

**Table S4.** Subdistribution hazard ratios and diagnostics from Fine–Gray competing risks regression models for cancer mortality and incident cancer.

| | | Cancer Mortality (N = 374, Events = 9) | | | | | | Cancer Incidence (N = 350, Events = 31) | | | | | |
|---|---|---|---|---|---|---|---|---|---|---|---|---|---|
| **Clock Category** | **Predictor** | **Beta** | **SHR (95% CI)** | **p-value** | **FDR p-value** | **ZPH** | **C-index** | **Beta** | **SHR (95% CI)** | **p-value** | **p-value** | **ZPH** | **C-index** |
| **Systemic** | Conventional | 0.38 | 1.46 (0.74–2.89) | 0.273 | 0.505 | 0.26 | 0.87 | -0.04 | 0.96 (0.61–1.50) | 0.849 | 0.986 | 0.63 | 0.68 |
| | PAC | 0.03 | 1.03 (0.97–1.10) | 0.345 | 0.505 | 0.30 | 0.87 | -0.02 | 0.98 (0.95–1.03) | 0.460 | 0.986 | 0.66 | 0.68 |
| | HPS | -1.74 | 0.18 (0.01–2.09) | 0.169 | 0.505 | 0.36 | 0.88 | 0.63 | 1.88 (0.30–11.98) | 0.504 | 0.986 | 0.53 | 0.68 |
| | ProtAge | 0.11 | 1.12 (0.89–1.40) | 0.341 | 0.505 | 0.12 | 0.87 | 0.13 | 1.14 (0.96–1.34) | 0.128 | 0.677 | 0.70 | 0.69 |
| **Organ-Specific** | Immune | 1.54 | **4.65 (1.91–11.27)** | **<0.001** | **0.015** | 0.70 | 0.90 | 0.27 | 1.32 (0.85–2.03) | 0.215 | 0.677 | 0.05 | 0.68 |
| | Liver | 0.62 | 1.86 (0.72–4.77) | 0.197 | 0.505 | 0.82 | 0.87 | 0.23 | 1.26 (0.80–1.99) | 0.315 | 0.866 | 0.63 | 0.68 |
| | Artery | 0.29 | 1.33 (0.57–3.14) | 0.512 | 0.626 | 0.62 | 0.87 | 0.15 | 1.17 (0.65–2.08) | 0.602 | 0.986 | 0.22 | 0.68 |
| | Intestine | 0.37 | 1.45 (0.71–2.96) | 0.302 | 0.505 | 0.65 | 0.87 | 0.09 | 1.10 (0.70–1.71) | 0.682 | 0.986 | 0.37 | 0.68 |
| | Adipose | 0.79 | 2.21 (0.46–10.63) | 0.323 | 0.505 | 0.24 | 0.89 | -0.03 | 0.97 (0.33–2.82) | 0.955 | 0.986 | 0.61 | 0.68 |
| | Kidney | 0.47 | 1.60 (0.86–3.00) | 0.140 | 0.505 | 0.64 | 0.88 | -0.04 | 0.96 (0.58–1.61) | 0.890 | 0.986 | 0.03 | 0.68 |
| | Pancreas | 0.65 | 1.92 (0.87–4.22) | 0.104 | 0.505 | 0.92 | 0.88 | 0.39 | 1.48 (0.87–2.51) | 0.150 | 0.677 | 0.08 | 0.69 |
| | Skin | 0.37 | 1.45 (0.71–2.98) | 0.310 | 0.505 | 0.15 | 0.87 | 0.18 | 1.20 (0.48–3.00) | 0.702 | 0.986 | 0.22 | 0.68 |
| | Brain | 0.38 | 1.47 (0.62–3.46) | 0.383 | 0.527 | 0.70 | 0.87 | -0.01 | 0.99 (0.64–1.53) | 0.981 | 0.986 | 0.59 | 0.68 |
| | Stomach | 0.15 | 1.16 (0.28–4.81) | 0.835 | 0.938 | 0.32 | 0.88 | 0.12 | 1.12 (0.41–3.11) | 0.823 | 0.986 | 0.20 | 0.68 |
| | Esophagus | 1.28 | 3.61 (0.28–46.79) | 0.326 | 0.505 | 0.51 | 0.88 | 0.56 | 1.74 (0.44–6.90) | 0.428 | 0.986 | 0.97 | 0.68 |
| | Adrenal | 3.03 | 20.71 (0.40–1069.55) | 0.132 | 0.505 | 0.95 | 0.87 | -2.15 | 0.12 (0.01–2.55) | 0.172 | 0.677 | 0.18 | 0.70 |
| | Muscle | 0.65 | 1.91 (0.70–5.23) | 0.209 | 0.505 | 0.84 | 0.87 | 0.06 | 1.07 (0.53–2.16) | 0.857 | 0.986 | 0.18 | 0.68 |
| | Heart | 0.05 | 1.06 (0.28–4.04) | 0.938 | 0.938 | 0.15 | 0.87 | -0.49 | 0.61 (0.29–1.28) | 0.191 | 0.677 | 0.87 | 0.69 |
| | Pituitary | -0.62 | 0.54 (0.12–2.41) | 0.420 | 0.543 | 0.66 | 0.86 | -0.35 | 0.70 (0.22–2.29) | 0.559 | 0.986 | 0.38 | 0.68 |
| | Salivary | -0.05 | 0.95 (0.34–2.67) | 0.921 | 0.938 | 0.98 | 0.87 | -0.61 | 0.54 (0.26–1.12) | 0.100 | 0.677 | 0.23 | 0.71 |
| | Thyroid | —* | —* | —* | —* | 0.19 | 0.87 | —* | —* | —* | —* | 0.47 | 0.68 |
| | Lung | 1.26 | 3.53 (0.90–13.82) | 0.070 | 0.505 | 0.92 | 0.90 | 0.65 | 1.92 (0.84–4.42) | 0.124 | 0.677 | 0.22 | 0.69 |

Subdistribution hazard ratios (SHRs) and 95% confidence intervals (CIs) correspond to a 1-SD increment in each standardized proteomic clock estimate derived from multivariable Fine–Gray subdistribution hazard models. Both models were adjusted for lifestyle covariates (age, sex, body mass index, smoking status, and alcohol consumption). For cancer mortality (N = 374, 9 cancer deaths), non-cancer mortality was treated as the competing risk event. For cancer incidence (N = 350, 31 incident cases), all-cause mortality prior to cancer diagnosis was treated as the competing risk event. Sample sizes reflect complete-case data for covariates and competing-risk follow-up; event numbers are identical to primary Cox models (9 deaths, 31 cases). Bold values indicate statistical significance following correction for multiple testing across all 22 clocks using the Benjamini–Hochberg false discovery rate (FDR $p < 0.05$). Proportional subdistribution hazards assumptions were evaluated using Schoenfeld-type residuals (ZPH), where $p > 0.05$ indicates no violation of the assumption. Model discrimination was evaluated using Harrell's concordance index (C-index). *Estimates for the Thyroid clock are not reported (—) due to extreme variance inflation resulting from limited event counts. *Beta, regression coefficient; CI, confidence interval; C-index, Harrell's concordance index; FDR, false discovery rate; HPS, Healthspan Proteomic Score; PAC, Proteomic Age Clock; SD, standard deviation; SHR, subdistribution hazard ratio; ZPH, test of proportional hazards based on Schoenfeld residuals.*
